# Infoveillance to Analyze Covid19 Impact on Central America Population

**DOI:** 10.1101/2020.05.26.20113514

**Authors:** Josimar Edinson Chire Saire, Roselyn Lemus-Martin

## Abstract

Infoveillance is an application within the Infodemiology field with the aim of monitoring public health and create public policies. Latin American countries have a different context about economics and health, so Infoveillance can be a useful tool to monitor and improve the decisions and be more strategical during the COVID-19 pandemic. The aim of this paper is to illustrate how data generated through Twitter can be used to help the implementation of strategies to address pandemic emergence in countries with Spanish as a native language in Central America by using a Text Mining Approach with Twitter as a data source in the capital of those countries.

## I. Introduction

The large-scale spread of COVID-19 [2] has constituted a huge impact on individuals, the economy of countries and overall in having to adapt to a new manner of living our daily life. Due to the nature of this pandemic, there is a huge need for new approaches to collect and process data in order to detect and manage the outbreaks faster [14] since large quantities of data are being produced by national and international government agencies, hospitals and clinicians.

The infodemiology or infoveillance approach in public health is based on Internet Based Sources (IBSs) and is particularly beneficial when outbreaks of a new disease appear since it can provide crucial information for locations where there is a limited capacity for traditional public health surveillance to detect both an acute outbreak or a changes in a trend in order to be able to give pandemic control recommendations [6].

Due to the nature of the COVID-19 epidemic, there is a crucial need for a fast data collection and posterior processing [10]. Large volumes of COVID-19 data are continuously gathered and analysed by various laboratories, health providers, and government agencies at national and international levels. The data gathering in traditional surveillance systems is very costly and slow with periods of months and weeks between the occurrence of an event and its report [13]. In the contrary, the infodemiology approach provides health agencies with the capability of providing a rapid response in order to reduce the mortality due to a pandemic or epidemic.

Information on the Internet related to health constitutes an opportunity to improve conventional public health surveillance during epidemics by providing opportune recognition of infectious diseases and a more direct access to data [17]. The importance of social media and information generated by Internet users is explicit, specially in a field that depends on defined reporting communication systems between the healthcare personnel, the laboratories and the governmental offices.

During a pandemic, acquiring information in real time is critical.The world has experienced previous pandemic of emerging diseases that require easy access to knowledge in order to manage the outbreaks and patients in an effective manner [5]. One big example was the Ebola epidemic, when it appeared, the world was connecting on Twitter to learn about this disease in real time [9]

Twitter is a social media platform where almost half a billion discuss diverse topics including those related to health. It has proven useful for different applications in public health ranging from geolocation to monitoring outbreaks and health emergency situations [7]

In 2011, Twitter was referred as an “ essential tool for every physician” [1] since it constitutes a forum for physicians to communicate in real time with each other to discuss the latest practices which is so important for the advancement of the medical field.

Twitter is so important in the dissemination of infectious disease news during a pandemic or epidemic. It played an important role as one of the most frequent resources used to learn about the H1N1 pandemic [8]. Signorini et al. [16] demonstrated the utility of Twitter in the dissemination of information on H1N1 and its influence on improving the vaccination rates [15].

Twitter has not only proven useful in the H1N1 pandemic outbreaks but also in the surveillance of other infectious diseases such as the MERS [4], Zika [12], Chikungunya [11] and Measless [3].

Because there is little scientific literature of studies reporting analysis on how the dissemination of the new coronavirus is affecting different social aspects in the American continent, we proposed a text mining approach in this paper to analyze the impact of COVID-19 in the Central American region.

## II. Methods

The present research uses techniques from Natural Language Processing and Data Mining to explore and analyze data per country. Next subsections present the details of the different experiments performed.

### A. Parameters

- date: 02-05-2020 to 09-05-2020
- terms: covid19, coronavirus
- geolocalization:
- language: Spanish
- radius: around 50 km

### B. Preprocessing

The data can contain symbols, number, links and more, so a cleaning process is necessary to have a standardized text. First, we transformed to lowercase, then eliminated characters(i.e. /, @ –,;) and finally we deleted the stop-words, which are words with no useful meaning i.e. articles, in/on, etc.

### C. Visualization

Data contain fields such as date and text. We started the analysis by plotting the frequency per day. After some classic questions are formulated, common words per day after some questions can be plotted. Examples of the formulated questions are: “ what is the term most relevant on X day? or who is the user with more posts?, who are these users?”

## III. Results

### A. Number of posts per capital?

The first question to answer is detecting how posts are produced in the capitals of each country. Figure 1 presents Costa Rica as the country with the highest number of posts and Honduras and Nicaragua as the countries with the lowest numbers.

**Fig. 1.**
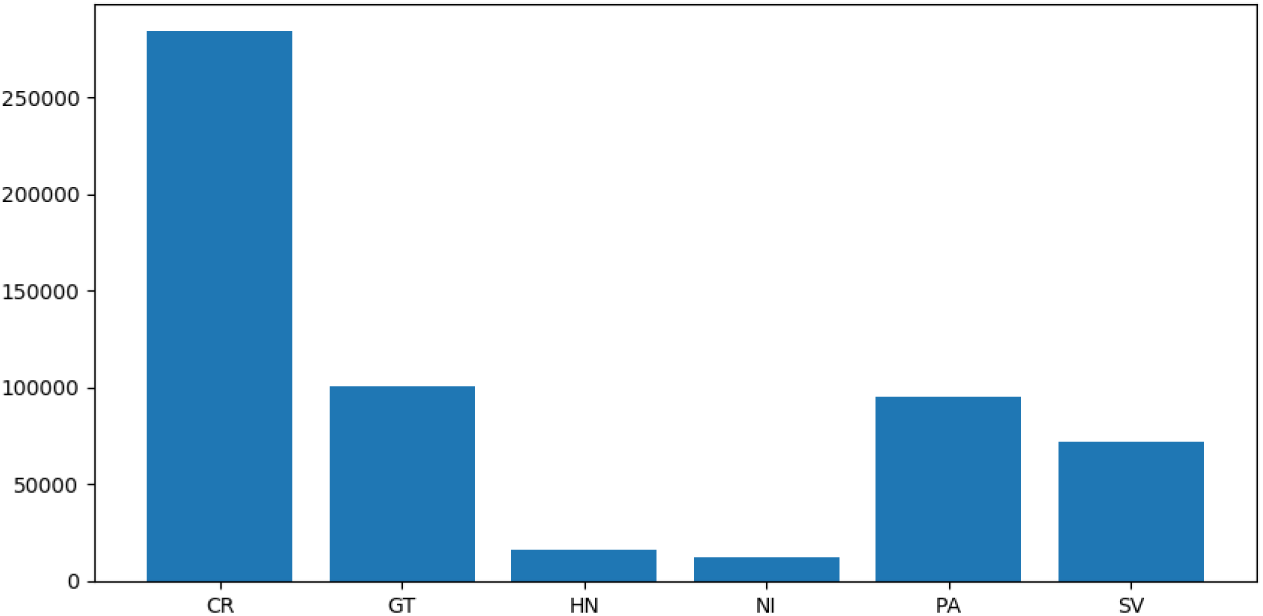
Number of posts per country

One derived question is how the frequency of posts per day in every country is. Most of the countries kept a steady trend for days with no major changes, except for Honduras that showed a significant peak on April 7th, so we can assume that there was an event of a surge in cases causing this trend change, see Fig. 2.

**Fig. 2.**
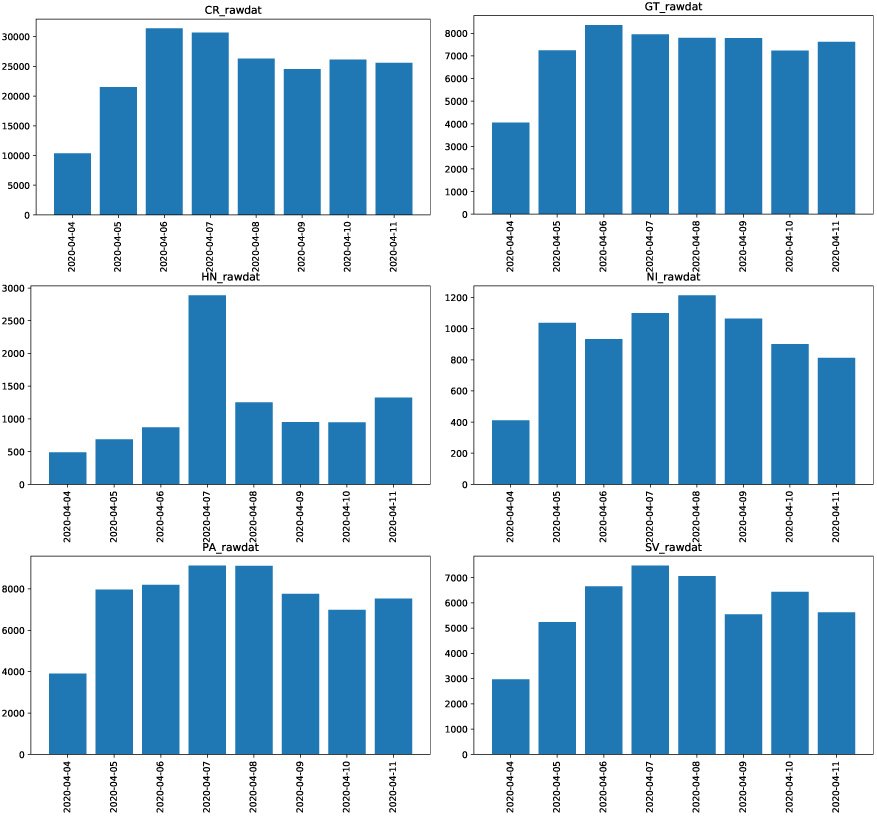
Number of posts per day: a) Costa Rica, b) Guatemala, c) Honduras, d) Nicaragua, e) Panama, f) El Salvador

### B. How are the top users in every country?

We consider only the top 100 of users d to find terms or n-grams with *n* = 2.

In Fig. 3 it can be observed that the top 5 of users were mainly wrriten or TV media for Costa Rica, Nicaragua, Panama and El Salvador. For Guatemala y Costa Rica individual accounts and media constitute the top 5 of users

**Fig. 3.**
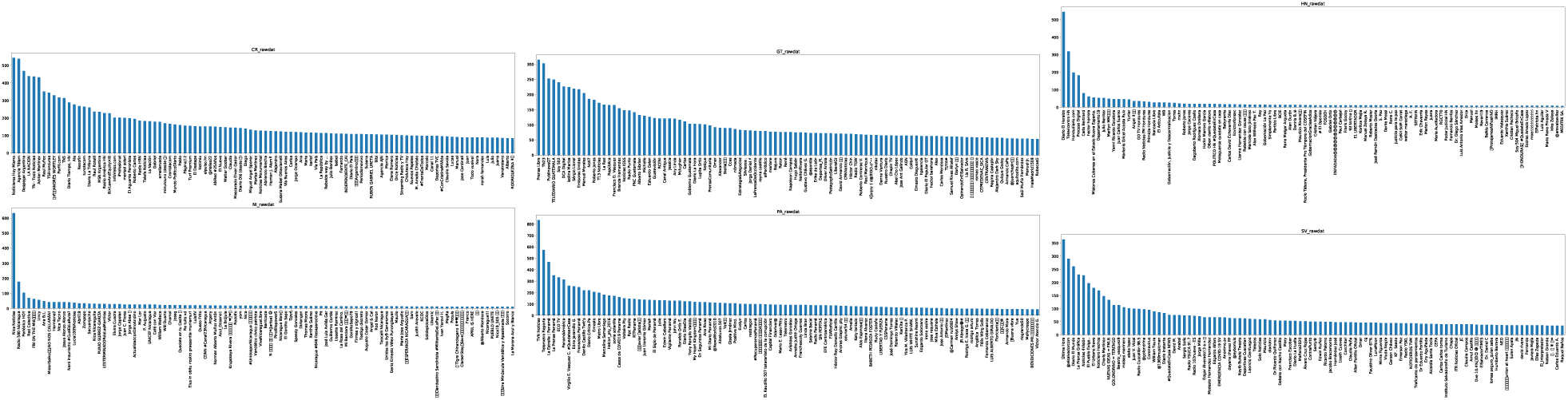
Top 100 users: a) Costa Rica, b) Guatemala, c) Honduras, d) Nicaragua, e) Panama, f) El Salva

### C. What is people talking about?

Considering image Fig.4, the next highlights are found:

**Fig. 4.**
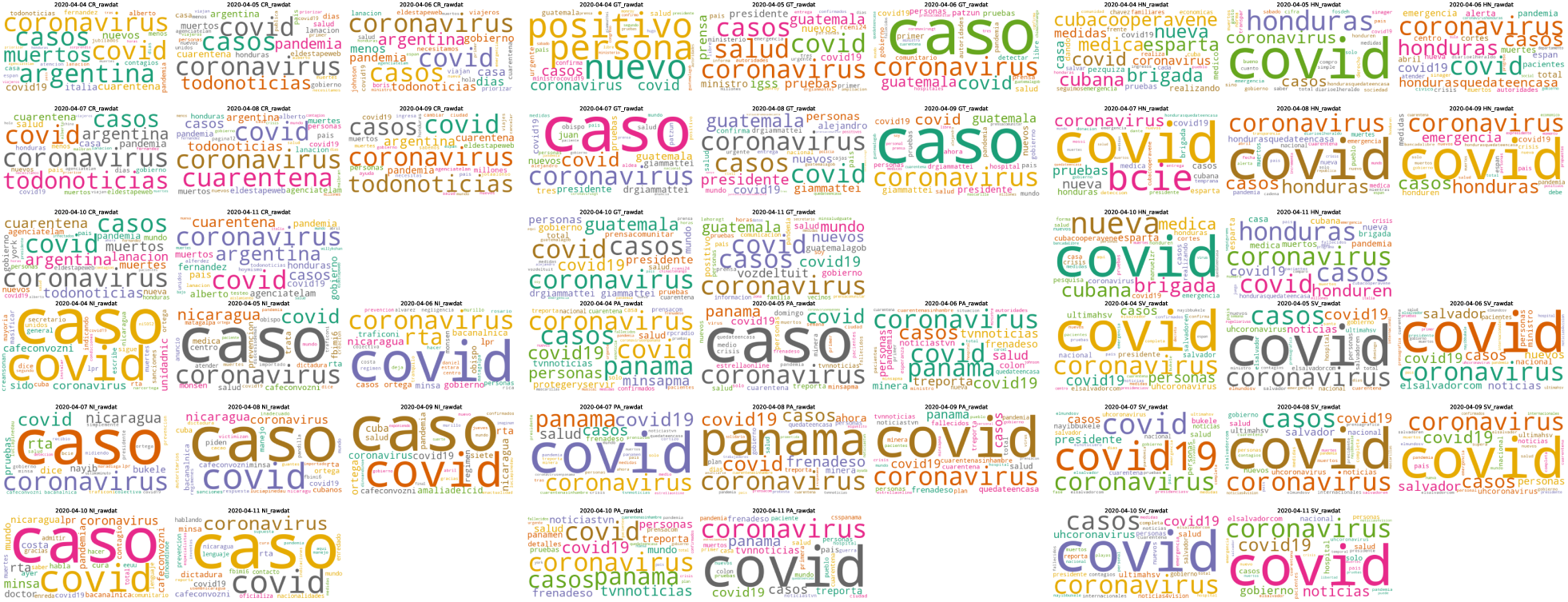
Cloud of words: a) Costa Rica, b) Guatemala, c) Honduras, d) Nicaragua, e) Panama, f) El Salvador

- The countries are talking about COVID-19, coronavirus and national cases from Costa Rica, to El Salvador.
- Words to express the actual context in every country are in Spanish and can be different in writing but posses similar meaning.
- Particularly, Costa Rica has the term ‘Argentina’ on consecutive days, so this can imply that people are observing and talking about what it is happening on Argentina and Honduras has a similar behaviour with ‘Cuba’.
- Only Costa Rica has ‘cuarentena’ as a term meaning quarantine during all the week from April 4th to April 11th and an increasing weight of the term on April 8th.

Next figure Fig.5 presents the next findings:

**Fig. 5.**
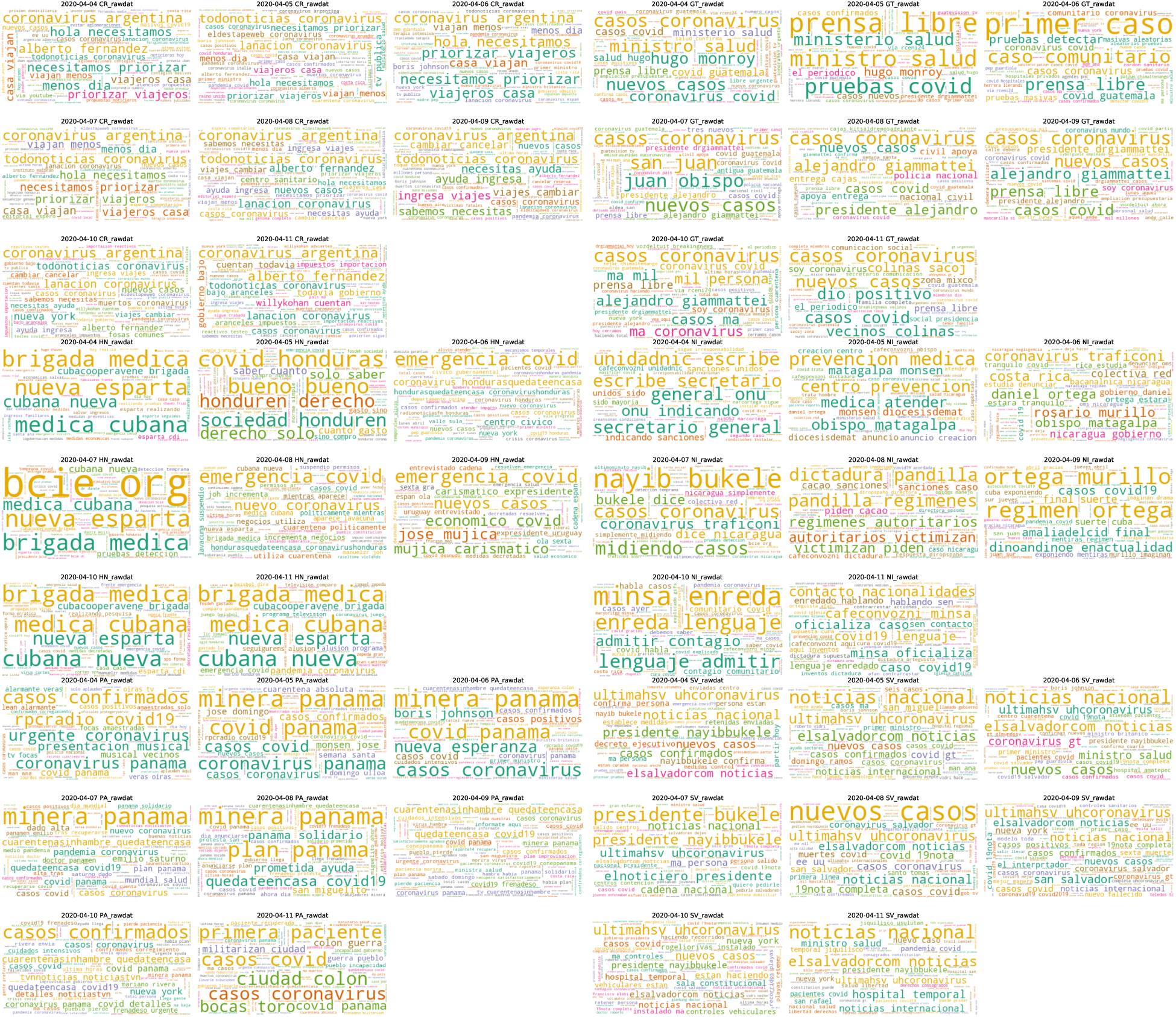
Cloud of wordbis: a) Costa Rica, b) Guatemala, c) Honduras, d) Nicaragua, e) Panama, f) El Salvador

- Costa Rica shows bigrams such as: ‘viajeros casa’, ‘priorizar pasajeros’, ‘ingresa viajes’ and these bigrams express a constant concern among people from Costa Rica who are stranded in other countries.
- On April 6th, Guatemala presents the term ‘primer caso’ with the highest frequency so it means that people are talking about the first detected case in this country.
- Term ‘emergencia covid’ on April 6th,8 th and 9th express a constant worry about COVID-19 emergency and ‘brigada medica’ on April 4th,10th and 11th is related to medical concerns.
- By contrast, Nicaragua has the term ‘prevention medica’ then people are talking about prevention, being this a different scenario not present in the other countries in the region.
- Panama presents the terms ‘casos covid’, ‘casos coronavirus’, ‘casos confirmados’ expressing cases of covid19 on this population.
- El Salvador has ‘casos nuevos’ during the week and higher number of hits of ‘nuevos casos’ on April 8th.

## IV. Conclusions

In this research paper, we present a Text mining approach using techniques from Natural Language Processing and Data Mining to explore and analyze Twitter data in the capitals of the main countries in the Central American Region.

This article shows the importance of Twitter usage in countries where the traditional epidemiological surveillance and monitoring of rural or non accessible areas can be very difficult.

Our study exhibits the fact that countries who are last in the ranking for internet and social media usage in Latin America such as Honduras and Nicaragua can benefit from social media searches as a tool to monitor the surge of cases in each country.

The experimental techniques were focused on analyzing the most searched terms on Twitter for the period between April 4th and April 11th providing interesting information such as the number of posts per capital, who the main users in every country are and most importantly what the most searched terms were. As we can conclude from our study, this surveillance tool is useful both for detection of surge in cases and for prevention in the case of Guatemala and Nicaragua.

## Data Availability

The data will be released on the github repository

